# Evaluating metabolome-wide causal effects on risk for psychiatric and neurodegenerative disorders

**DOI:** 10.1101/2024.11.06.24316481

**Authors:** Lachlan Gilchrist, Julian Mutz, Pirro Hysi, Cristina Legido-Quigley, Sulev Koks, Cathryn M. Lewis, Petroula Proitsi

## Abstract

Evidence indicates phenotypic and biological overlap between psychiatric and neurodegenerative disorders. Further identification of underlying mutual and unique biological mechanisms may yield novel multi-disorder and disorder-specific therapeutic targets. The metabolome represents an important domain for target identification as metabolites play critical roles in modulating a diverse range of biological processes. Here, we used Mendelian randomisation (MR) to test the causal effects of ∼1000 plasma metabolites and ∼300 metabolite ratios on anxiety, bipolar disorder, depression, schizophrenia, amyotrophic lateral sclerosis, Alzheimer’s disease, Parkinson’s disease and multiple sclerosis. In total, 85 causal effects involving 77 unique metabolites passed FDR correction and robust sensitivity analyses (IVW-MR OR range: 0.73-1.48; *p_FDR_* < 0.05). No evidence of reverse causality was identified. Multivariate analyses implicated sphingolipid metabolism in psychiatric disorder risk and carnitine derivatives in risk for amyotrophic lateral sclerosis and multiple sclerosis. However, polygenic risk scores for prioritised metabolites showed limited prediction in the UK Biobank. Downstream colocalisation in regions containing influential variants identified greater than suggestive evidence (PP.H4 ≥ 0.6) for a shared causal variant for 29 metabolite/psychiatric disorder trait-pairs on chromosome 11 at the *FADS* gene cluster. Most of these metabolites were lipids containing linoleic or arachidonic acid. Additional colocalisation was identified between the ratio of histidine-to-glutamine, glutamine, Alzheimer’s disease and *SPRYD4* gene expression on chromosome 12. Although no single metabolite had a causal effect on a psychiatric and a neurodegenerative disease, results suggest a broad effect of lipids across brain disorders. Metabolites identified here may help inform future targeted interventions.

## 1. Introduction

Psychiatric disorders show substantial co-morbidity^1^ and shared genetic architecture^2^. Additionally, they are common both prior to and within neurodegenerative diseases^3–7^ – themselves genetically related^8,9^ – and are indicated as risk factors for their later onset^10–12^. Genomic analyses suggest that psychiatric and neurodegenerative risk are partially underpinned by shared risk loci and causal brain transcripts and proteins^13–17^. By further investigating shared biological risk mechanisms, novel interventions may be identified with effectiveness for specific or multiple disorders.

Metabolomics – the study of small molecules linked to metabolism – is a promising tool for such investigation as it represents the stage of the omics cascade closest to the phenotype^18^. Importantly, metabolites are strong drug target candidates as many are obtained dietarily and modulate diverse biological processes including gene expression, RNA metabolism and protein activity^19–21^. Obtaining samples is minimally invasive as thousands of metabolites can be detected in plasma, serum or urine^22^.

Similar metabolite groups, such as ceramides, are indicated to play a role in both psychiatric and neurodegenerative disorders^23–27^. However, observational studies may be confounded by reverse causation^28^. Importantly for psychiatric and neurodegenerative disorders, the metabolome can be perturbed by disease pathology and/or pharmaceutical interventions^29–33^. As such, following onset and treatment discerning the direction of effect and identifying causal metabolite targets is challenging. These limitations can be addressed using Mendelian randomisation (MR), a statistical framework that leverages genetic variants as instrumental variables (IVs) to proxy an exposure and infer its causal effect on an outcome of interest^34^. As risk alleles are randomly assorted and fixed from conception, MR is less vulnerable to reverse causality than classical epidemiological approaches^28,35^. Beneficially, this approach can be performed using summary statistics from large genome-wide association studies (GWAS) (two-sample MR (2SMR)), allowing researchers to perform increasingly powerful analyses^36^. This approach applied to the metabolome has already identified several causal risk metabolites for Alzheimer’s disease, bipolar disorder, major depressive disorder, and multiple sclerosis^37–41^.

However, no study has yet been conducted to identify shared and unique metabolomic risk factors across the psychiatric and neurogenerative disorder spectrum. In this study, we assess the causal effect of ∼1000 unique plasma metabolites and ∼300 metabolite ratios on four psychiatric disorders (anxiety, bipolar disorder, depression and schizophrenia) and four neurodegenerative diseases (amyotrophic lateral sclerosis (ALS), Alzheimer’s disease (AD), Parkinson’s disease (PD) and multiple sclerosis (MS)). We further prioritise key metabolites using Bayesian model averaging, identify regions of shared causal variants with statistical colocalisation – integrating expression quantitative trait loci (eQTLs) – and assess the predictive performance of polygenic scores for risk metabolites on incident diseases in the UK Biobank (UKB).

## 2. Methods

An overview of the analyses is shown in **Figure 1**.

**Figure 1:**
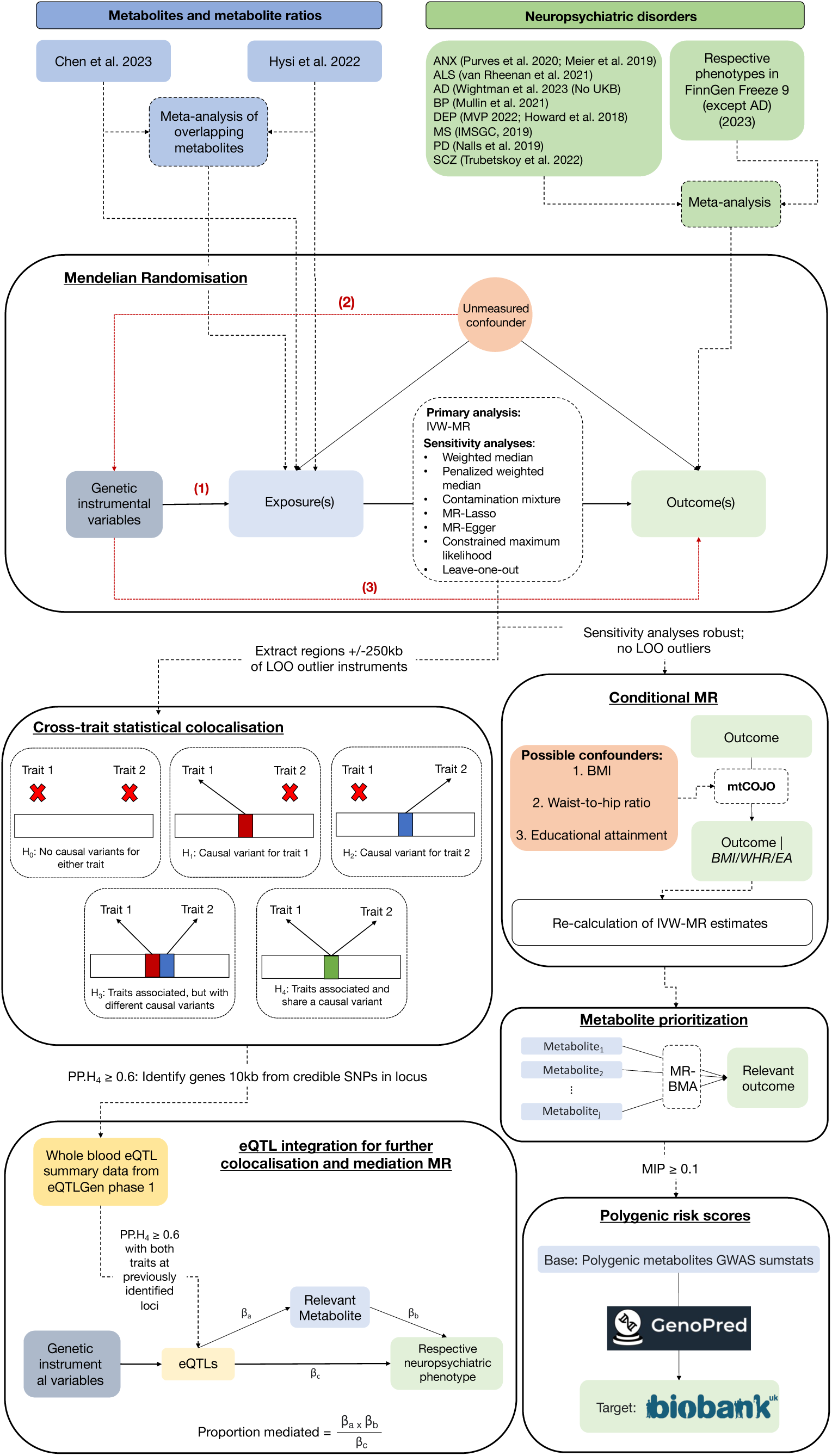
Analysis flowchart. In the Mendelian randomisation section, the three key assumptions are illustrated and can be described as: (1) Instruments must be robustly associated with the exposure – usually defined as genome-wide statistical significance of the variants; (2) Instruments are not associated with a confounder and (3) Instruments are not associated with the outcome via pathways other than through the exposure, for example through horizontal pleiotropy. These assumptions are known as the (1) relevance, (2) independence and (3) exclusion restriction assumption, respectively. ANX = anxiety; ALS = amyotrophic lateral sclerosis; AD = Alzheimer’s disase; BP = bipolar disorder; DEP = depression; MS = multiple sclerosis; PD = Parkinson’s disease; SCZ = schizophrenia

### 2.1 GWAS summary statistics

#### 2.1.1 Neuropsychiatric disorders

An overview of the GWAS summary statistics is shown in **Table 1**. These consisted of individuals of European ancestry only. For each disorder, we performed inverse variance weighted (IVW) genome-wide meta-analysis across samples^9,42–51^ using METAL^52^ to maximise sample size and statistical power. Details on each sample can been seen in the original papers. For AD, we used summary statistics that did not contain family history-based proxy phenotyping from the UK Biobank given its noted influence on the effect direction of downstream analyses^53^. The *APOE* region (chr19:45,020,859–45,844,508 (GRCh37)) was excluded from the AD GWAS due to known pleiotropic effects on relevant non-AD diseases such as heart disease and hypercholesterolaemia^54^. It was not meta-analysed with FinnGen freeze 9 as with the others as the sample already contained samples from a previous FinnGen freeze. Genetic correlations between these disorders – calculated using Linkage Disequilibrium Score Regression (LDSC)^55^ in GenomicSEM^56^ v.0.0.5 – are described and plotted in **Supplementary Material 1** and **2**, respectively. Full output tables are in **Supplementary Tables 1** and **2**. Effective sample sizes were calculated as per Grotzinger et al.^57^, providing comparable sample size estimates representing equivalently powered GWAS with a 1:1 case/control ratio.

**Table 1:**
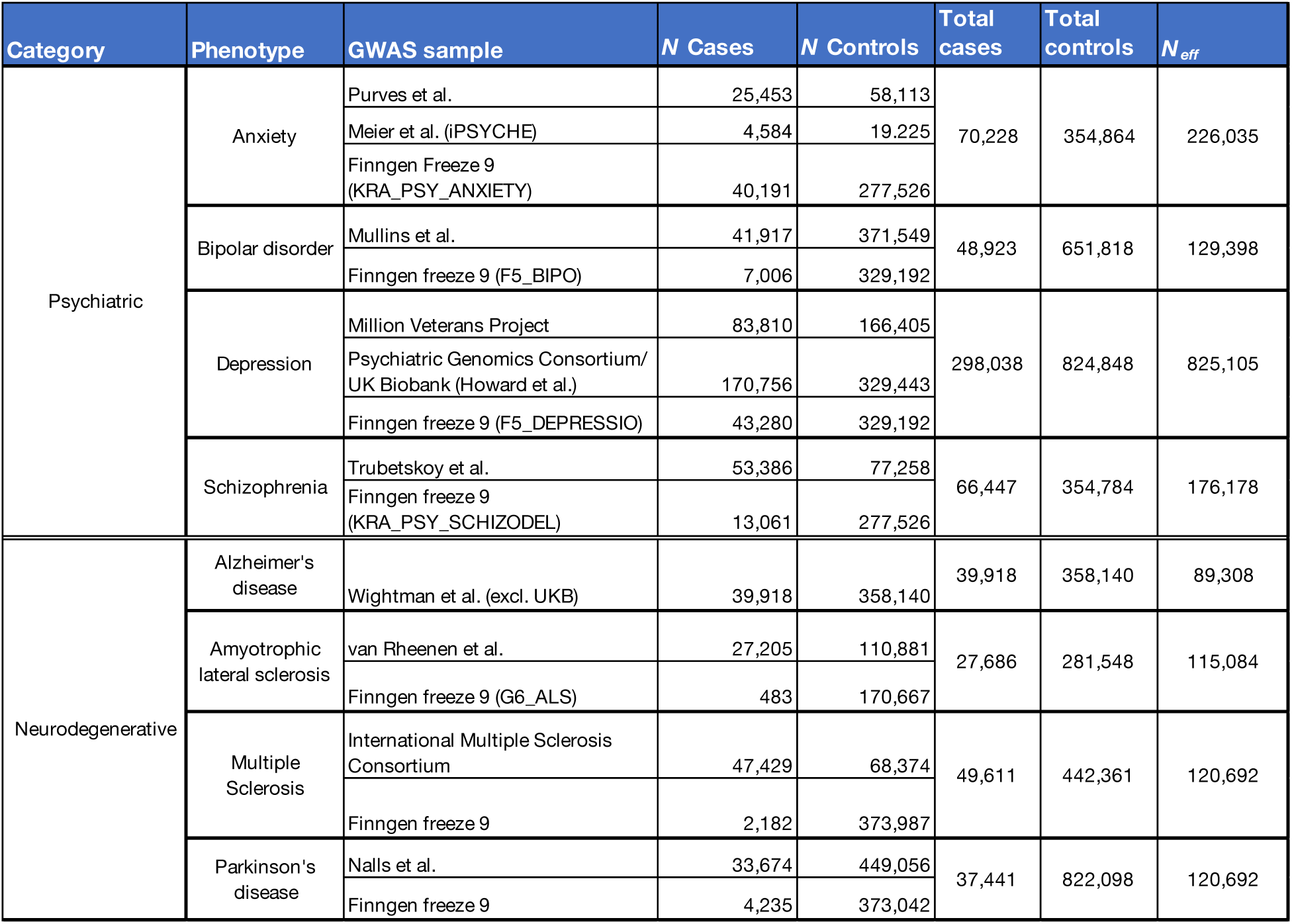
Previously conducted genome-wide association study summary statistics for psychiatric and neurodegenerative disorders used in this study.

#### 2.1.2 Plasma metabolites

We obtained metabolite summary statistics from two studies conducted in non-overlapping samples: (1) 1,091 individual metabolites and 309 metabolite ratios measured in 8,299 unrelated individuals from the Canadian Longitudinal Study of Aging (CLSA)^58^ and (2) 722 individual metabolites measured in 8,809 individuals in the NIHR UK Bioresource cohort^59^. In both studies, metabolites were detected and quantified in plasma by Metabolon Inc. using ultra-high performance liquid chromatography-tandem mass spectroscopy. Unidentified metabolites were excluded from analyses. We performed genome-wide meta-analysis as above for 431 metabolites when the same metabolite was available in both samples for a meta-analysed sample size of 17,038.

#### 2.1.3 GWAS standardisation

Prior to meta-analysis, summary statistics were standardised with MungeSumstats^60^ version in R (v.4.2.1) using dbsnp 144 and the BSgenome.Hsapiens.1000genomes.hs37d5 reference genome available through Bioconducter (v.3.1.3). Missing rsids were obtained, duplicate and multi-allelic variants excluded, and alleles aligned to the reference, flipping effect and frequency direction accordingly. Where necessary, MungeSumstats was used to lift over genome build coordinates from GRCh38 to GRCh37 via the UCSC Genome Browser chain file.

### 2.2 Mendelian randomisation

#### 2.2.1. Metabolite instrument selection

Independent instrumental variables (IVs) were initially selected at genome-wide significance (*p* ≤ 5 x 10^-8^), clumping at an r^2^ threshold of 0.001 within 10,000 kb of the lead variants. Clumping was conducted using the ieugwasr package (v.0.1.4), PLINK (v.1.9)^61^ binaries and the European sample of the 1000 Genomes phase 3 reference panel^62^ (*N* = 503), restricted to minor allele frequency (MAF) > 0.01. This is the same reference panel used by the MRC IEU OpenGWAS database^63^ (http://fileserve.mrcieu.ac.uk/ld/1kg.v3.tgz).

Where less than five IVs were available for any given exposure, a lower *p*-value threshold of 5 x 10^-6^ was used for IV selection as per previous analyses^64^. If less than 5 IVs were available at the 5 x 10^-6^ threshold, this metabolite was excluded. Where an IV was not available for the outcome, proxy IVs (i.e., variants in linkage disequilibrium (LD) with the original instrument at r^2^ > 0.8) were identified using snappy (v.1.0) (https://gitlab.com/richards-lab/vince.forgetta/snappy) and same reference panel. Proxies were selected using the highest r^2^ value and closest genomic position. Prior to analysis, exposure and outcome were harmonised to the same effect allele, and strand ambiguous palindromic variants (MAF > 0.42) dropped. Instrument strength was assessed via their F-statistic (β_exposure_^2^/S_Eexposure_^2^), with weak instruments (F-statistic < 10) excluded. If less than 5 IVs remained, the metabolite was again excluded. Instrument measurement error was assessed with the I^2^G-X statistic (< 0.9 suggestive of measurement error)^65^.

Following the above steps the effects of 428 meta-analysed metabolites were tested, plus 740 and 132 metabolites from the CLSA and NIHR UK Bioresource studies, respectively, for a total of 1300 unique metabolites/metabolite ratios.

#### 2.2.2. MR analyses

We conducted the primary analyses using inverse variance weighted MR (IVW-MR) with multiplicative random effects as recommended^66^. If the IVW-MR estimate was statistically significant after Benjamini-Hochberg false discovery rate (FDR) correction^67^ (*p_FDR_* ≤ 0.05, corrected for total number of tests *specific to each* outcome), six sensitivity analyses were conducted to assess its robustness to violations of the pleiotropy assumption and instrument validity. We implemented methods that assume a majority valid instruments (weighted median^68^; penalised weighted median^69^), plurality valid instruments (constrained maximum likelihood (cML)^70^; contamination mixture model MR (MR-ContMix)^71^), and that exclude invalid outliers (MR-Lasso^72^). Additionally, we used MR-Egger^73^, which gives consistent causal estimates even if all instruments are not valid, provided pleiotropic effects are not correlated with variant-exposure associations. This is known as the Instrument Strength Independent of Direct Effect (InSIDE) assumption and is MR-Egger-specific. To pass sensitivity criteria, all sensitivity analysis point estimates needed to be directionally concordant with the IVW estimate and the majority (≥ 4) statistically significant at *p* ≤ 0.05. Where a metabolite passed sensitivity criteria, we performed reverse IVW-MR as above with the outcome as the exposure.

Pleiotropy was assessed using the MR-Egger intercept test (no evidence of pleiotropy: *p_Egger-Intercept_* ≥ 0.05) and heterogeneity using Cochran’s Q (no evidence of heterogeneity: *p_Q-Stat_* ≥ 0.05). Metabolites passing sensitivity were further inspected using leave-one-out (LOO) analysis to assess whether causal estimates were driven by the inclusion of a single influential variant. Metabolites that passed sensitivity criteria and had no LOO outlier variant were considered polygenic metabolites and are referred to as such henceforth. Those passing sensitivity but with a LOO outlier variant were considered single instrument metabolites.

### 2.2 Further analysis of polygenic metabolites

#### 2.3.1 Conditional analysis

Hysi et al.^59^ noted that many metabolites are genetically correlated with phenotypes related to body composition and educational level. To assess whether any identified effects were driven in part by the genetic correlations between the metabolites and these phenotypes, we performed conditional GWAS of the neuropsychiatric disorders using multi-trait conditional and joint analysis (mtCOJO)^74^ to remove the per-SNP effects of body mass index (BMI), BMI-adjusted waist-to-hip ratio (WHR), and educational attainment (EA).

We obtained GWAS summary statistics for BMI (*N* = 806,834) and BMI-adjusted waist-to-hip ratio (*N* = 697,734) from the GIANT consortium^75,76^. We additionally obtained summary statistics for educational attainment (EA) from the EA4 GWAS meta-analysis, excluding 23andMe, from the Social Science Genetic Association Consortium (SSGAC)^77^ (*N* = 765,283).

IVW-MR estimates were recalculated for significant metabolite-neuropsychiatric disorder pairs post-mtCOJO and compared to the original estimates. Forest plots were visually inspected for evidence of attenuation between original and adjusted estimates.

#### 2.3.2 Metabolite prioritisation using MR-BMA

Metabolites are highly genetically correlated^58^. Consequently, univariate MR estimates – which assume exposures are independent – may be biased by the pleiotropic effects of other risk metabolites. Such issues may be addressed using MR with Bayesian model averaging (MR-BMA)^78^. MR-BMA is a multivariable MR approach that ranks of causal effects among related exposures according to their independent causal signal. Unlike conventional multivariable MR methods, MR-BMA is well suited for the highly correlated nature of high-throughput exposures such as metabolites^78^. Instruments were selected by pooling those used in IVW-MR for all per-disorder statistically significant metabolites, excluding correlated instruments as before (r^2^ > 0.001, window = 10,000 kb). We use a prior probability of 0.1 and a prior variance of 0.25 as previously^78,79^. Metabolites were ranked according to their marginal posterior probability (MIP), with MIP ≥ 0.1 interpreted as causal. To avoid multicollinearity between the exposures, we calculated a genetic correlation matrix using the linkage disequilibrium score regression (LDSC)^80^ function of GenomicSEM^56^. As per the original MR-BMA study, where point estimates ≥ 0.95, we excluded the metabolite correlated with the most other metabolites (if > one) or retained only the metabolite with the most-significant IVW-MR estimate in the MR-BMA analysis (if only one).

#### 2.3.2 Cross-trait polygenic score (PGS) analysis

To assess whether any of the polygenic metabolites prioritised in MR-BMA be used for disease prediction, we calculated polygenic scores (PGS) – the weighted sum of an individual’s risk alleles^81^ – within the reference standardised framework of the GenoPred pipeline^82^ using individual level data from the UK Biobank (UKB)^83^. Within GenoPred, allele weights were calculated using MegaPRS^84^, a PGS method that uses the BLD-LDAK heritability model and improves prediction by incorporating information on allele frequency, LD and various functional annotations. MegaPRS has shown strong predictive performance compared to other PGS methods^85,86^.

UKB participants were excluded from analyses if they had unusual levels of heterozygosity, a call missing rate >2% or discordant phenotypic and genetic sex (F_X_ < 0.9 for males, F_X_ > 0.5 for females). Individuals were filtered for relatedness up to 3^rd^ degree relatives based on KING kinship estimates (r < 0.044)^87^ using GreedyRelate (v1.2.1) to remove one individual from each related pair, maximising sample size (https://gitlab.com/choishingwan/GreedyRelated). Analyses were restricted to individuals of European ancestry. The final sample contained 381,564 individuals prior to any age-based exclusions.

Case/control status was defined using the first occurrence data fields from the UKB (**Supplementary Table 3**), where cases were ascertained using primary care Read v2 or Read CTV3 codes, hospital inpatient ICD-9 and ICD-10 codes, death record ICD-10 codes and medical condition codes self-reported at UKB assessment centre visits. Individuals were excluded if the code had no event date or if the date of event was before, the same year as or on the participants date of birth, or a date in the future. Associations between metabolite-PGS and respective neuropsychiatric disease was assessed using logistic regression, controlling for age, sex, genotyping batch, assessment centre and the first ten genetic principal components from the UKB. Within each model, the comparison group was restricted to individuals with no occurrence of any of the neuropsychiatric disorders. To provide age-appropriate controls for AD and PD, samples were restricted for those analyses to individuals aged ≥60 and ≥50 years, respectively. For DEP and ANX cases, individuals with an occurrence of BP or SCZ were excluded. The variance explained by each PGS was assessed using Nagelkerke’s pseudo-r^2^. Sample sizes ranged from 140,154 to 356,763, depending on the disorder, with case numbers ranging from 600 (ALS) to 46,900 (DEP) (mean age range = 56.65-64.13; %male range = 46.32-49.35).

### 2.4 Further analysis of single instrument metabolites

#### 2.4.1 Statistical colocalisation

Using LOO analysis, we identified IVs that significantly attenuated the IVW-MR estimate when excluded. We extracted regions +/- 250 kb of these IVs and performed statistical colocalisation for each metabolite and the respective neuropsychiatric disorder within the COLOC-reporter pipeline^88^. Regions of interest were extracted and harmonised for each phenotype to contain only shared variants on the reference panel. Colocalisation was performed using the *coloc.abf* function of the COLOC package^89^. A posterior probability of 0.8 for hypothesis four (PP.H_4_) – presence of a shared causal variant between the phenotypes – was taken as evidence of colocalisation (PP.H4 ≥ 0.6 taken as suggestive).

#### 2.4.2 eQTL colocalisation

For regions of colocalisation (including suggestive), we identified genes located within 10 kb of credible SNPs using Ensembl and biomaRt (v.2.54.0). For each gene, we extracted *cis-*eQTLs from the whole blood eQTL summary data of the eQTLGen Consortium^90^ (*N* = 31,684) and performed colocalisation between the eQTLs and both phenotypes in each metabolite-neuropsychiatric disorder pair as above.

#### 2.4.2 eQTL MR

For genes colocalising with both the metabolite and neuropsychiatric disorder a trait-pair, we extracted independent instruments for gene expression from the eQTL summary statistics (*p* ≤ 5 x 10^-8^; r^2^ ≤ 0.001; window = 10,000 kb) and used IVW-MR to assess the causal effect of gene expression levels on the outcome and exposure. We examined the consistency of the gene expression causal effects using Summary-based Mendelian Randomisation (SMR)^91^, extracting eQTL probes for the relevant gene from blood-based eQTL summary datasets from Westra et al.^92^ (*N* = 3,511) and Lloyd-Jones et al.^93^ (*N* = 2,765). Heterogeneity was assessed using the HEterogeneity In Dependent Instruments (HEIDI) test, with p_HEIDI_ < 0.05 indicative of heterogeneity. This is essentially a test of colocalisation.

Previous research has indicated that the causal effect of gene expression on a complex trait can be mediated by the effect of metabolites within a transcript-metabolite-trait causal triplet^94^. Where gene expression levels showed a significant causal effect on both an outcome and a metabolite, we calculated the proportion mediated using the product of coefficients method as per previous analyses^95^. Standard errors were estimated using the delta method and *p*-values drawn from a z-score distribution.

## 3. Results

### 3.1 Mendelian randomisation

We conducted a total of 10,327 tests of the causal effect of the metabolites and ratios on the eight neuropsychiatric outcomes (instrument *N*-range: 5-27). Of these, 1695 tests were conducted with instruments clumped at 5 x 10^-8^ and 8632 with instruments clumped at 5 x 10^-6^. For single instruments, F-statistics ranged from 10.11 to 5625.88 (mean per-test F-statistics range: 20.77-1163.90). Analyses were thus not impacted by weak instruments. Per-test I^2^G-X ranged from 0.45 to 1.000. Only 94 tests (0.009%), involving 13 unique metabolites, were conducted with I^2^G-X < 0.9, suggestive of measurement error. Only 27 tests were conducted using instruments with I^2^G-X ≤ 0.8, involving six metabolites: 2-hydroxyarachidate, 4-hydroxy-2-oxoglutaric acid, 4-vinylguaiacol sulphate, glycolithocholate, and O-cresol sulphate.

Primary IVW-MR analyses identified 138 causal effects involving 113 unique metabolites after outcome-specific FDR correction (**Supplementary Table 4**; **Figure 2a**). Of these, 85 metabolite-outcome pairs passed our sensitivity criteria, involving 77 unique metabolites (**Supplementary Table 5**). All neuropsychiatric disorders had two or more metabolites with statistically significantly effects. No reverse effects were detected for these metabolite-outcome pairs after FDR-correction (**Supplementary Table 6**). IVW-MR odds ratios (OR) for metabolite-neuropsychiatric disorder trait-pairs ranged from 0.73 to 1.48 (*p*-value range: 1.52 x 10^-3^ - 9.67 x 10^-31^; *p_FDR_* range: 4.45 x 10^-2^ - 1.26 x 10^-27^). However, MR-Egger intercept tests detected significant pleiotropy for eight of the 85 metabolite-outcome pairs (*p_Egger-Intercept_* < 0.05) (**Supplementary Table 7**). Cochran’s Q heterogeneity tests detected significant heterogeneity for 15 (*p_Q-Stat_* < 0.05) (**Supplementary Table 8**). All metabolite-outcome pairs with *pEgger-Intercept* < 0.05 and 14 out of 15 with *p_Q-Stat_* < 0.05 were identified as influenced by single influential variants in the LOO analysis (**Supplementary Table 9**). Only the effect of the ratio of mannose to glycerol on depression (*p_Q-Stat_* = 0.043) was not driven by a single influential variant.

**Figure 2:**
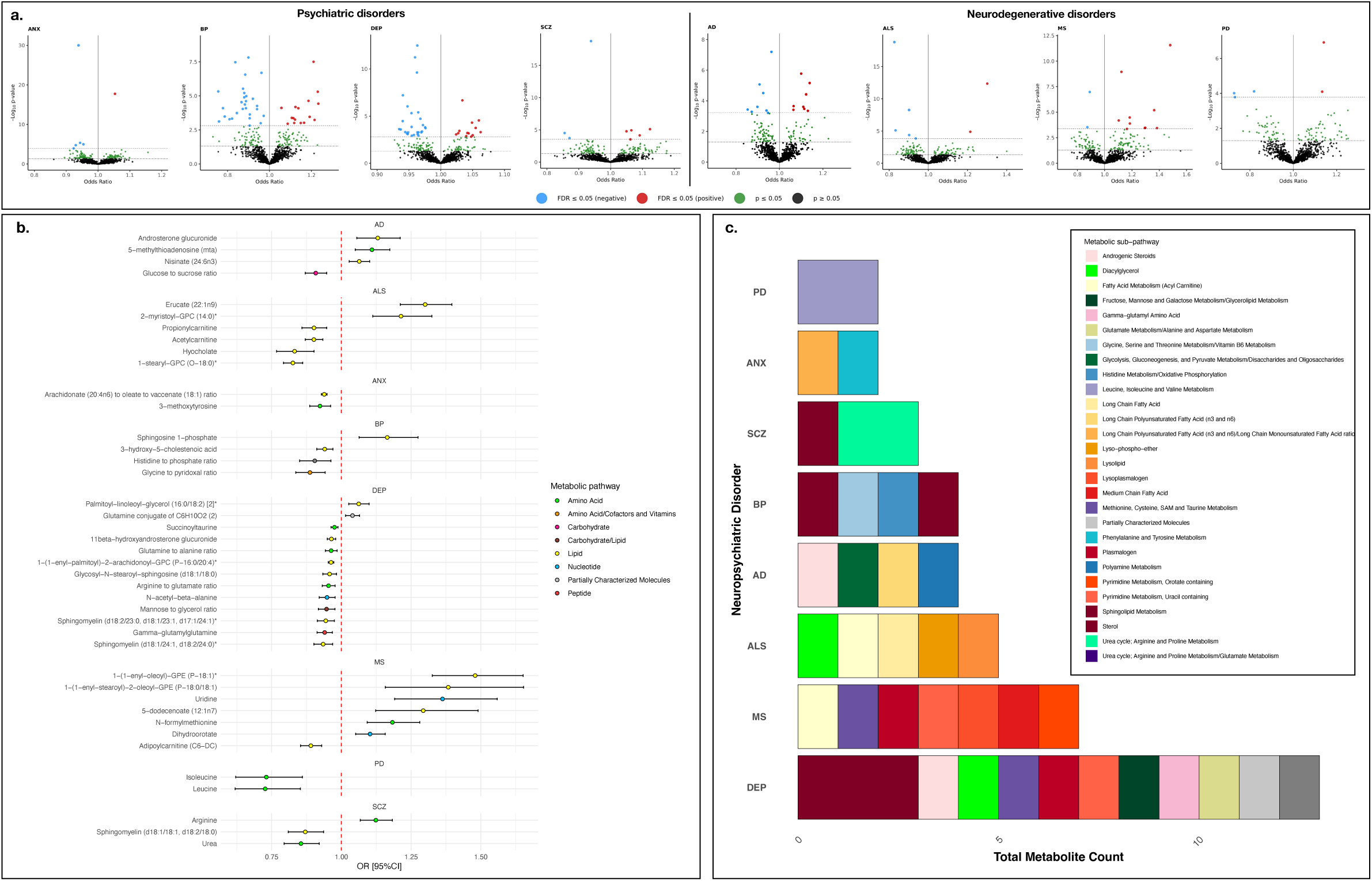
(a). Volcano plots for anxiety (ANX), bipolar disorder (BP), depression (DEP), schizophrenia (SCZ), Alzheimer’s disease (AD), amyotrophic lateral sclerosis (ALS), multiple sclerosis (MS) and Parkinson’s disease (PD) showing odds ratios and -log10 p-values from inverse variance weighted Mendelian randomisation (IVW-MR) for all tested metabolites. (b). Forest plot of FDR statistically significant metabolites associated with neuropsychiatric outcomes in IVW-MR and passing sensitivity tests, with no leave-one-out (LOO) outliers (i.e., polygenic metabolites). Point estimates indicate odds ratios with bars representing 95% confidence intervals (95%CI). Point estimates for each metabolite are coloured according to their metabolic super-pathway. (c). Stacked histograms showing total count of FDR statistically significant polygenic metabolites associated with each neuropsychiatric disorder, with fill colours indicating metabolite sub-pathways. AD = Alzheimer’s disease; ALS = Amytropic Lateral Sclerosis; ANX = anxiety; BP = Bipolar Disorder; DEP = Depression; MS = Multiple Sclerosis; PD = Parkinson’s disease; SCZ = Schizophrenia

Excluding metabolites with effect estimates primarily influenced by single variants left a total of 41 metabolite-neuropsychiatric disorder trait-pairs with polygenic metabolites – all unique. Of these, 13 were identified with depression, seven with MS, six with ALS, four each with AD and bipolar disorder, three with schizophrenia, and two each with anxiety and PD (**Figure 2b**). These metabolites were linked to nine broad metabolic classes (super-pathways), counting the combinations of pathways of metabolite ratios as unique pathways (**Figure 2b**; **Supplementary Table 10**). Over half (*N* = 22) were lipids, and the rest amino acids (*N* = 10), nucleotides (*N* = 3), peptides (*N* = 1), carbohydrates (*N* = 1) or ratios involving amino acid/cofactors and vitamins (*N* = 1), amino acid/energy (*N* = 1) and carbohydrate/lipid (*N* = 1) pathways. The remaining metabolite – glutamine conjugate of C6H10O2 (2) – is considered only partially characterised.

These metabolites were implicated in 28 unique sub-pathways representing the metabolic/biochemical subclasses related to the metabolites (**Figure 2c**; **Supplementary Table 10**). The largest group of metabolites was related to sphingolipid metabolism (*N* = 5). All metabolites involved in sphingolipid metabolism had effects on psychiatric disorder risk. Sphingosine 1-phosphate has a risk increasing effect on bipolar disorder (OR [95%CI] = 1.17 [1.06-1.28], *p*-value = 9.49 x 10^-4^, *p_FDR_* = 0.03), while glycosyl-N-stearoyl-sphingosine (d18:1/18:0), sphingomyelin (d18:1/24:1, d18:2/24:0) and sphingomyelin (d18:2/23:0, d18:1/23:1 and d17:1/24:1) has risk decreasing effects on depression (OR range: 0.93-0.96, *p*-value range: 2.38 x 10^-4^ – 9.91 x 10^-4^; *p_FDR_* range: 0.02-0.04). Additionally, sphingomyelin (d18:1/18:1, d18:2/18:0) has a risk decreasing effect on schizophrenia (OR [95%CI] = 0.87 [0.81-0.94], *p*-value = 1.89 x 10^-4^, *p_FDR_* = 0.04).

The strongest negative effect was observed for the amino acid leucine and PD (OR [95%CI] = 0.73 [0.62-0.85], *p* = 9.64 x 10^-5^; *p_FDR_* = 3.13 x 10^-2^). Isoleucine showed similar effects (OR = 0.73 [0.62-0.86]; *p* = 1.65 x 10^-4^; *p_FDR_* = 4.28 x 10^-2^). The strongest positive effect was observed for the lipid 1-(1-enyl-oleoyl)-GPE (P-18:1) and MS (OR = 1.48 [1.33-1.65]; *p*-value = 2.71 x 10^-12^; *p_FDR_* = 3.42 x 10^-9^). The most statistically significant effect was identified for the ratio of arachidonate (20:4n6) to oleate to vaccenate (18:1) and anxiety (OR = 0.93 [0.93-0.95], *p*-value = 9.67 x 10^-31^, *p_FDR_* = 1.26 x 10^-27^).

The remaining 44 metabolite-neuropsychiatric disorder trait-pairs had attenuated effects in the LOO analysis, involving 36 unique metabolites and six of the neuropsychiatric outcomes (**Supplementary Table 11**). Most of the effects were observed in relation to psychiatric disorders (*N* = 40), primarily with BP (*N* = 16) and DEP (*N* = 16). Another four were observed with ANX and three with SCZ. These metabolite-psychiatric trait-pairs involved 33 unique metabolites, 25 of which were lipids and one a ratio of lipid/carbohydrate. These lipids were involved in 12 unique sub-pathways, the largest group of which was related to phospholipid metabolism (*N* = 6). The remainder were amino acids (*N* = 5), a carbohydrate (*N* = 1) and a xenobiotic (*N* = 1), involved in seven unique sub-pathways. Of those remaining, four unique single instrument metabolites had a causal effect AD – two amino acids and two xenobiotics – and a nucleotide showed causal effect on MS.

A total of 31 of the LOO associations were driven by 11 influential variants on chromosome 11 within a ∼500 kb window (61293499-61854782 bp). This region contains the fatty acid desaturase (*FADS*) gene cluster, which is linked to the regulation of fatty acid levels and their circulation^96^. All metabolites influenced by these variants were lipids and associated with psychiatric disorder risk (*NANX* = 4, *NBP* = 13, *NDEP* = 12 and *NSCZ* = 2). Between-variant LD estimates for the influential instruments in this region were calculated using LDlinkR (v.1.3)^97^ and ranged from 0.16-1, with r^2^ ≥ 0.72 for all variants except rs14570 and rs2524299 which showed a maximum r^2^ of 0.34 and 0.38, respectively (**Figure 3a**; **Supplementary Table 12**).

**Figure 3:**
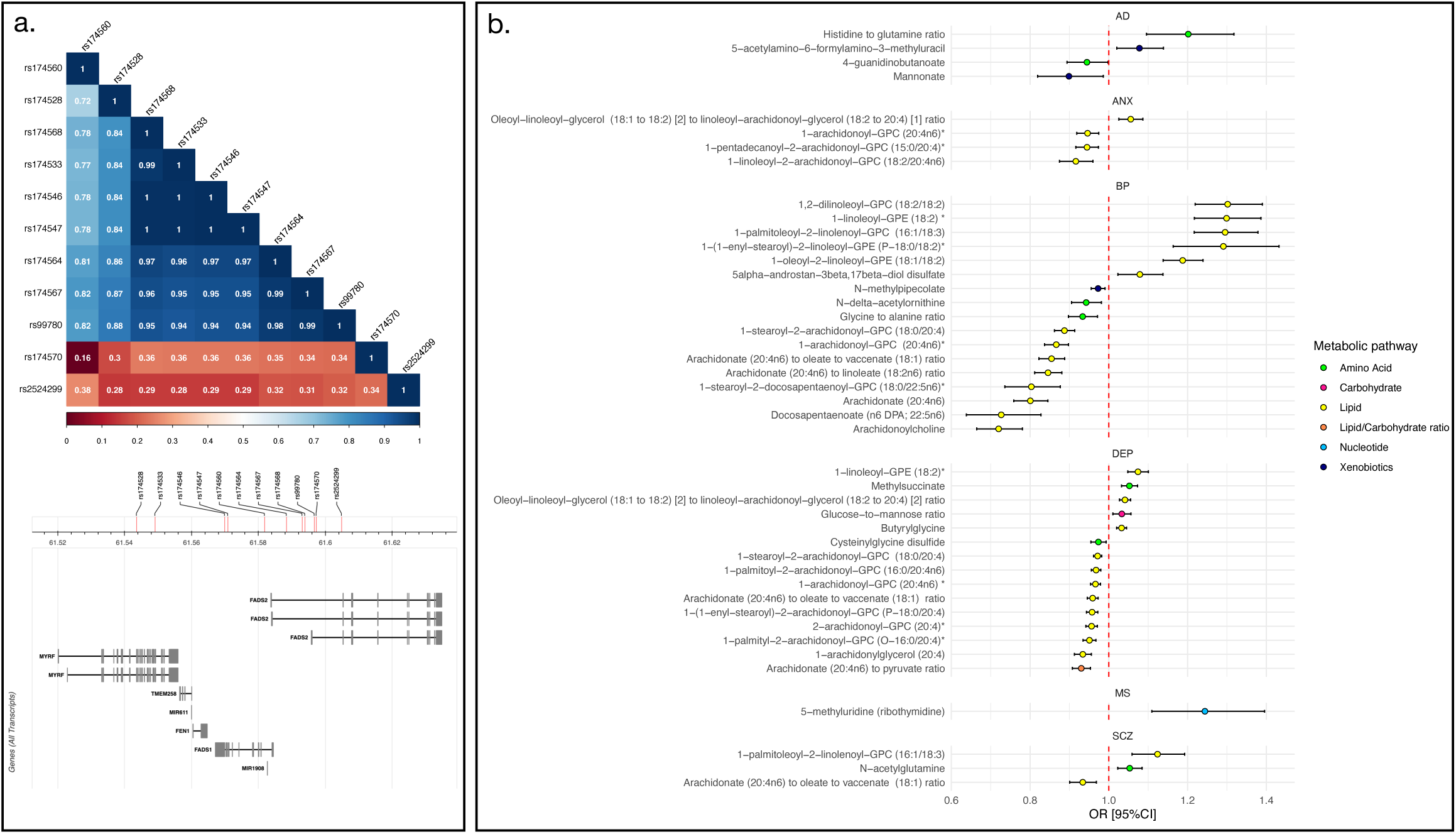
(a). The r^2^ estimates between influential single variants on chromosome 11 alongside their genomic location. (b). Recalculated Wald ratio estimates for metabolites with influential single variants using the influential variant only. Point estimates indicate odds ratios with bars representing 95% confidence intervals (95%CI). Point estimates for each metabolite are coloured according to their metabolic super-pathway. AD = Alzheimer’s disease; ANX = anxiety; BP = Bipolar Disorder; DEP = Depression; MS = Multiple Sclerosis; SCZ = Schizophrenia

We used Wald ratio tests to re-estimate the effect of single instrument metabolites on the relevant outcome using the influential instrument only. F-statistics ranged from 105.37 to 5625.88. Wald estimates were directionally consistent with IVW-MR estimates and remained significant at *p* ≤ 0.05, although the protective effects of 4-guanidinobutanoate and mannonate on AD were only nominally significant (*p* = 0.04 and 0.02, respectively) (**Figure 3b**; **Supplementary Table 13**). Regions containing influential instruments were further investigated using colocalisation.

### 3.2 Further analysis of polygenic metabolites

#### 3.2.1 Conditional analysis

Conditioning neuropsychiatric outcomes on BMI, BMI-adjusted WHR or EA did not attenuate the IVW-MR effect estimates for causally associated metabolites when re-estimated (**Supplementary Table 15**; **Supplementary Material 3**). This suggests that the metabolite associations were not influenced by any correlation between the genetic instrument’s effect and the effects of these traits as captured in the neuropsychiatric GWAS.

#### 3.2.2 Metabolite prioritisation using MR-BMA

Prior to the per-outcome multivariate MR modelling in MR-BMA, we excluded the less statistically significant of any metabolite pair correlated ≥ 95% (**Supplementary Table 15**). As such, we excluded 5-methylthioadenosine (mta) from the AD model due to its correlation with the glucose to sucrose ratio, 5-dodecenoate (12:1n7) from the MS model due to its correlation with N-formylmethionine and uridine, 2-myristoyl-GPC (14:0)* from the ALS model due to its correlation with 1-stearyl-GPC (O-18:0)* and the glutamine to alanine ratio from the DEP model due to its correlation with gamma-glutamylglutamine.

For AD, anxiety, MS, PD and schizophrenia, all metabolites had MIPs ≥ 0.1, suggesting unique causal contributions to risk. For bipolar disorder, sphingosine 1-phosphate, the ratio of histidine to phosphate and the ratio of glycine to pyridoxal passed the MIP threshold, with 3-hydroxy-5-cholestenoic acid just under (MIP = 0.098). However, of the 12 metabolites included for depression, only the two sphingomyelins (d18:1/24:1, d18:2/24:0 and d18:2/23:0, d18:1/23:1, d17:1/24:1) and 1-(1-enyl-palmitoyl)-2-arachidonoyl-GPC (P-16:0/20:4) passed. For ALS, only acetylcarnitine passed the MIP threshold. Acetylcarnitine had the highest overall MIP of all metabolites tested (MIP = 0.893) (**Supplementary Table 16**).

#### 3.2.3 Polygenic score (PGS) analysis

No MR-BMA prioritised metabolite PGS was associated with any of their respective outcomes after FDR correction for 23 tests (*p_FDR_* ≥ 0.05). Nominal associations were observed between depression and the PGS for sphingomyelin (d18:2/23:0, d18:1/23:1, d17:1/24:1) and 1-(1-enyl-palmitoyl)-2-arachidonoyl-GPC (P-16:0/20:4) with identical effect sizes (OR[95%CI] = 0.989 [0.979-0.998], *p*-value range = 2.131 x 10^-2^ - 2.517 x 10^-2^) and between the PGS for leucine and PD (OR [95%CI] = 0.962 [0.93-0.996, *p*-value = 2.814 x 10^-2^). These effects were directionally consistent with the MR estimates but had non-relevant predictive ability given small Nagelkerke’s pseudo-r^2^ estimates (Nagelkerke’s pseudo-r^2^MAX = 1.445 x 10^-4^) (**Supplementary Table 17**).

### 3.3 Further analysis of single instrument metabolites

#### 3.3.1 Colocalisation at loci of influential instruments

Of the 44 colocalisation tests conducted in regions +/-250 kb of the LOO influential variants, 26 showed evidence of colocalisation (PP.H4 ≥ 0.8) and a further five had suggestive evidence (PP.H4 ≥ 0.6) (**Supplementary Table 18**). All except one of these regions were on chromosome 11 within a ∼500 kb window (61293499-61854782 bp). All disorders colocalising with metabolites in this region were psychiatric and every psychiatric disorder examined showed colocalisation with at least one metabolite. Colocalisation has been previously observed between the omega-3 fatty acid docosahexaenoic acid (DHA) and depression within the same region^39^.

All implicated metabolites except 1-stearoyl-2-docosapentaenoyl-GPC (18:0/22:5n6)* were complex fatty acid chains containing the fatty acids arachidonic acid (AA) (*N* = 18), linoleic acid (LA) (*N* = 5), alpha-linolenic acid (ALA) (*N* = 1) or both AA and LA (*N* = 4). Higher levels of AA-containing metabolites were consistently protective in Wald estimates while higher levels of LA- or ALA-containing metabolites had risk increasing effects. Where ratios contained both AA- and LA-containing metabolites, higher levels of the AA relative to the LA was protective and higher LA metabolites relative to the AA risk increasing (**Supplementary Table 19**). This is concordant with previous metabolome-wide MR analysis of bipolar disorder^41^ and observational analysis of depression^98^. The other region of colocalisation was identified on chromosome 12 (56615338-57115338) between the ratio of histidine-to-glutamine and AD (PP.H4 = 0.92).

Two tests showed evidence of two distinct causal variants at the tested locus – the ratio of glucose to mannose and depression (chr2: 27480940-27980940; PP.H3 = 0.92) and docosapentaenoate (n6 DPA; 22:5n6) and bipolar disorder (chr11: 61347212-61847212; PP.H3 = 0.99). Suggestive evidence for two distinct causal variants was also identified for butyrylglycine and depression (chr12:120925524-121425524; PP.H3 = 0.70), methylsuccinate and depression (chr12:120926083-121426083; PP.H3 = 0.63), and 5-methyluridine (ribothymidine) and MS (Chr22:50687969-51187969; PP.H3 = 0.63). PP.H3 ≥ 0.6 in these regions suggests that MR estimated effects arose from linkage disequilibrium, violating MRs horizontal pleiotropy assumption. Remaining colocalisation tests were inconclusive.

#### 3.3.2 eQTL colocalisation

In total, nine genes were identified within 10 kb of credible SNPs at colocalising loci (**Supplementary Table 20**). For the colocalisation analyses at chr11:61293499-61854782, eQTLs for *MIR611* or *MIR1908* were not available in the eQTLGen summary statistics. As such, follow-up eQTL colocalisation in this region was conducted only with *MYRF*, *TMEM258*, *FEN1*, FADS2 and *FADS1*. For the region chr12:56615338-57115338, eQTLs were not available for *MIP*, but were available for *GLS2* and *SPRYD4*. In total, 130 colocalisation tests were conducted with eQTLs. Of these, only eQTLs for *SPRYD4* colocalised with both a risk metabolite and its respective neuropsychiatric outcome: the ratio of histidine-to-glutamine (PP.H4 = 0.91) and AD (PP.H4 = 0.91). Additional colocalisation was identified for *GLS2* with the ratio of histidine-to-glutamine (PP.H4 = 0.99), but not AD (PP.H4 = 0.42). *TMEM258* colocalised with depression (PP.H4 = 0.86). Suggestive colocalisation was identified between *TMEM258* and anxiety and between *MYRF* and depression (PP.H4 ≥ 0.6). Of the remaining tests, 113 indicated two distinct causal variants in the region (PP.H3 ≥ 0.8), four were suggestive of only a causal variant for gene expression, and the remainder inconclusive (**Supplementary Table 21**).

#### 3.3.3 eQTL MR for the histidine to glutamine ratio and AD

We conducted MR analyses for the effect of *SPRYD4* expression levels on the ratio of histidine-to-glutamine and AD using five independent instruments. *SPRYD4* expression levels were identified as increasing risk for AD (OR = 1.08 [1.02-1.16], β [95%CI] = 0.08 [0.02-0.15], *p*-value = 0.007) and the level of the ratio of histidine to glutamine (β [95%CI] = 0.30 [0.18-0.42], *p*-value = 1.16 x 10^-6^). SMR estimates for the effect of *SPRYD4* expression were directionally consistent and statistically significant at *p* ≤ 0.05 for AD and the ratio of histidine-to-glutamine across eQTL panels. HEIDI tests did not indicate heterogeneity (*pHEIDI* ≥ 0.05) (**Supplementary Table 22**). There was no evidence that the ratio of histidine to glutamine substantially mediates the effect of *SPRD4* expression on AD, with only a nominally significant association and wide confidence intervals (proportion mediated [95%CI] = 63.20% [1.80-124.59%], *p*-value = 0.043).

### 3.6 Further examination of histidine, glutamine, AD and *SPRYD4*

To further evaluate the association between the ratio of histidine to glutamine, we re-examined the IVW-MR estimates for the separate effects of histidine and glutamine on AD. No causal effect was observed for histidine on AD, not even nominally (OR = 1.11 [0.99-1.27], β = 0.11 [-0.011-0.24], *p*-value = 0.07). However, there was a nominally significant negative causal effect for glutamine (OR = 0.87 [0.81-0.94], β [95%CI] = -0.14 [-0.21 to -0.06], *p*-value = 0.0007, *p_FDR_* = 0.052). Although glutamine passed post-hoc sensitivity criteria with no evidence of pleiotropy or heterogeneity (**Supplementary Table 23**), LOO identified the same influential variant on chromosome 12 as observed for the ratio of histidine-to-glutamine – the 3′-UTR variant rs2657879 (**Supplementary Table 24**). For glutamine, the effect of rs2657879 was in the opposite direction to the histidine-to-glutamine ratio.

A Wald ratio test using only this instrument estimated a negative effect of glutamine on AD risk (OR = 0.84 [0.76-0.91], β = -0.19 [-0.28 to -0.09], *p*-value = 9.65 x 10^-5^). Strong evidence of colocalisation was observed between glutamine and AD in the region (PP.H4 = 0.92), and between glutamine and *SPRYD4* (PP.H4 = 0.98) (**Supplementary Table 25**). Gene expression levels of *SPRYD4* were negatively associated with glutamine in the MR analysis (β = -0.35 [-0.52 to -0.18], *p*-value = 3.94 x 10^-5^). Strong, directionally consistent effects of *SPRD4* expression on glutamine were observed in SMR (**Supplementary Table 26**). However, the HEIDI tests indicated heterogeneity (pHEIDI ≤ 0.05). There was no evidence that glutamine mediated the effects of SPRD4 expression on AD (proportion mediated [95%CI] = 75.81% [-0.36-151.99%], *p*-value = 0.051). LocusZoom plots for the ratio of histidine to glutamine, glutamine, AD and *SPRYD4* gene expression are shown in **Figure 4**.

**Figure 4:**
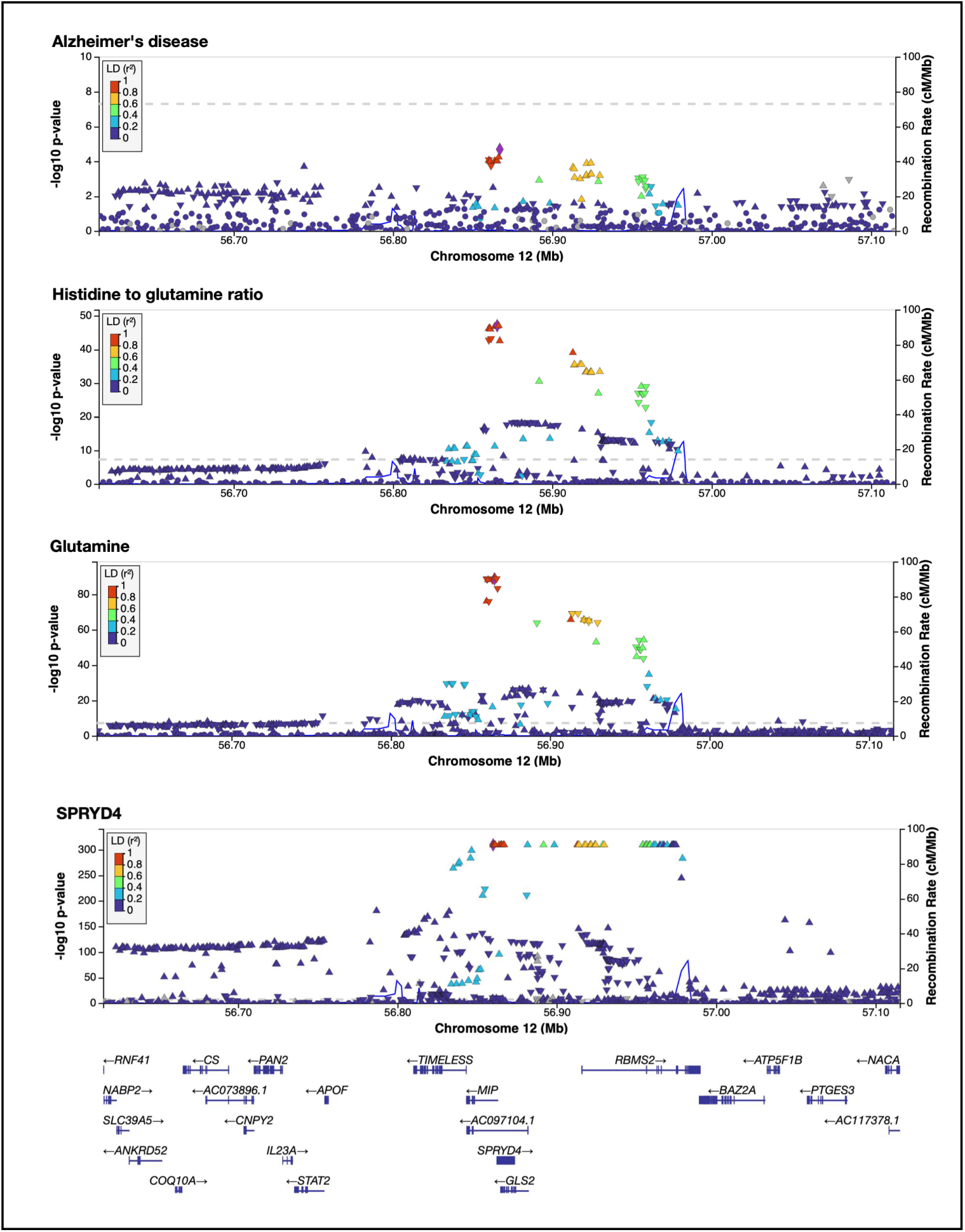
LocusZoom plots of the colocalising region (PPH4 ≥ 0.8) chr12: 56615338-57115338 for Alzheimer’s disease, the ratio of histidine to glutamine, glutamine, and SPRYD4 expression.

## 4. Discussion

We identified 77 unique plasma metabolites and a total of 85 sensitivity robust MR estimated causal effects on neuropsychiatric disorder risk. Over half of associations were driven by single influential variants. Of these, 30 metabolite-neuropsychiatric disorder trait-pairs had greater than suggestive colocalisation and 29 were lipids colocalising with a psychiatric disorder near the *FADS* gene cluster on chromosome 11. No eQTL colocalisation was observed for any metabolite-outcome trait-pair in this region, suggesting the underlying mechanisms are not related to shared gene expression effects. Additional colocalisation was observed between AD glutamine-related metabolites and *SPRYD4* gene expression on chromosome 12. For the remaining 41 polygenic metabolites, multivariate analysis with MR-BMA prioritised 23 metabolites and implicated sphingolipids and carnitine derivatives in psychiatric and neurodegenerative risk respectively. However, the predictive performance of metabolite polygenic scores was negligible in the UK Biobank.

Lipids – specifically polyunsaturated fatty acids (PUFAs) – have previously shown MR causal effects on depression, with docosahexaenoic acid (DHA) colocalising at the *FADS* gene cluster^39^. Our study highlights the broader significance of this region in linking lipid levels to risk across the psychiatric disorder spectrum. Here, we identify suggestive colocalisation between at least one lipid and every examined psychiatric disorder. Nearly all lipids colocalising with psychiatric disorders contained linoleic acid (LA) or arachidonic acid (AA) – both omega-6 PUFAs. AA-containing lipids were consistently protective, while LA-containing lipids increased risk. These findings are consistent with other MR studies of bipolar disorder, depression and schizophrenia^41,99,100^. Our results suggest that these associations extend to ANX. Although these effects were primarily driven by shared causal variants on chromosome 11, several polygenic metabolites comprised of AA/LA-containing lipids also showed causal effect on psychiatric risk. The ratio of arachidonate (20:4n6)-to-oleate-to-vaccenate (18:1) and 1-(1-enyl-palmitoyl)-2-arachidonoyl-GPC (P-16:0/20:4) – both AA-containing – had risk decreasing effects on anxiety and depression respectively and were prioritised in multivariate analysis. In single instrument analyses, this ratio had risk decreasing effects on bipolar disorder, depression and schizophrenia, colocalising with all. These results suggest that effects of AA/LA-containing lipids in psychiatric risk may extend beyond effects on chromosome 11.

Circulating levels of lipids containing LA and AA may be promising intervention targets for psychiatric disorders. LA – the most abundant omega-6 PUFA in the western diet – is found in nearly all manufactured food through vegetable oils^101^, while AA is found in animal products such as meat, poultry, fish and eggs^102^. As free fatty acids AA is a downstream product of LA, from which it is synthesised through desaturation and chain elongation^102^ (**Supplementary Material 4**).

Free LA and AA are linked to inflammatory dysregulation^103^ – itself implicated in psychiatric disorders^104,105^. AA is catalysed by cytochrome P450 enzymes to epoxyeicosatrienoic acids (EETs), which show anti-inflammatory effects and are suggested to be protective of neurological function^106^. The deleterious effects of LA-containing lipids compared to the protective effects of AA-containing lipids may implicate dysregulated conversion from LAs to AAs, contributing to elevated inflammation as less AA is available for conversion to anti-inflammatory EETs. However, LA-to-AA conversion is relatively low in humans and requires LA to be in free form following cleavage from lipid molecules via enzymes such as phospholipase A2 (PLA2). Although PLA2 action on fatty acids has been previously linked to inflammatory depression^107^, further investigation is required to elucidate mechanisms by which LA- and AA-containing lipids contribute to psychiatric risk.

The effects of the ratio of histidine-to-glutamine and glutamine on AD observed here are in line with previous MR evidence suggesting higher blood glutamine as protective^37,108^. Interestingly, glutamine is noted to exert a neuroprotective effect against β-amyloid aggregation^109^ – a key drug target for AD^110^. This study provides further evidence for its protective effects on AD risk and suggests the association is driven by shared causal variants on chromosome 12. Further, we observe that the association between AD and glutamine may be partially driven by the mutual effect of *SPRYD4* gene expression. As *SPRYD4* has not been previously identified in AD GWAS it represents a novel locus of interest for future study.

Our study also provides further evidence for the role of sphingolipid metabolism in psychiatric disorders. Sphingolipids are fatty acids with roles in cell growth/death, inflammation, mitochondrial function and immune response^111^. We identified two sphingomyelins and glycosyl-N-stearoyl-sphingosine (d18:1/18:0) as protective for depression, with the sphingomyelins both prioritised in multivariate analysis. Further, sphingomyelin (d18:1/18:1, d18:2/18:0) was protective for schizophrenia and sphingosine-1-phosphate indicated to elevate risk of bipolar disorder. Sphingomyelins are central to ceramide metabolism, to which they are converted via sphingomyelinase and back via sphingomyelin synthase^112^ (**Supplementary Material 5**). Ceramides are catabolised into sphingosine and eventually sphingosine-1-phosphate^112^. General dysfunction of sphingolipid metabolism, particularly ceramide aggregation, has been previously observed in psychiatric disorders^23,24,111,112^ and reduced sphingomyelin levels may contribute to elevated neuroinflammation^112^. As such, the ceramide system is a proposed pathway for drug-based interventions for psychiatric disorders^112^. This study implicates sphingomyelins as possible protective targets.

Although fewer metabolomic patterns were observed in neurodegenerative disorders we did observe protective effects of several carnitine related metabolites – adipoylcarnitine for MS and acetylcarnitine and propionylcarnitine for ALS. All three are metabolised from L-carnitine, which plays a role in the transportation of fatty acids to mitochondria^113^, and when acetylated is suggested to slow disease progression for both disorders and AD^114–116^. Further, lower serum carnitine levels are observed in ALS^117^ and are associated with increased fatigue in MS^118^. A randomised control trial (RCT) of acetylcarnitine supplementation in ALS suggests it may improve self-sufficiency in patients compared to placebo^114^. The importance of acetylcarnitine for ALS is further emphasised by our multivariate analysis, where it was the only metabolite prioritised for ALS and had the highest MIP of any metabolite examined. Interestingly, a larger phase two/three RCT of acetylcarnitine effects on biological and clinical outcomes in ALS is underway (https://classic.clinicaltrials.gov/ct2/show/NCT06126315). The remaining polygenic metabolites did not show clear functional relatedness, indicating disorder-specific metabolomic contributions – broadly confirmed in MR-BMA, where, for most disorders, all metabolites had a MIP ≥ 0.1, indicating unique causal effects.

Although no specific metabolites were identified with an effect on both a psychiatric and neurodegenerative disorder, our results suggest the general importance circulating lipids in both categories. Dietary lipids are known to modulate neuroinflammation and play an important role in the brain by regulating neuronal and synaptic function^119^. Future studies exploring biological overlap in neuropsychiatric disorders may therefore wish to focus on lipidomic analysis for the purposes of further elucidating shared metabolic pathways.

Several disorder-specific metabolites warrant further discussion. For example, we replicated the risk-increasing effect of uridine on MS previously identified observationally and in MR^40,120^. Uridine is linked to pyrimidine metabolism with important biological functions including RNA synthesis and the regulation of glucose, lipid and amino acid levels^121^. Interesting, within pyrimidine biosynthesis, uridine is an eventual metabolic product of dihydroorotate^121^, also identified here as increasing risk for MS. Additionally, the branch chain amino acids (BCAAs) leucine and its isomer isoleucine were both protective for PD. Plasma levels are noted to be lower in cases than controls and correlated with increased functional disability^122^. In an RCT of PD/parkinsonism, supplementation with leucine and vitamin D enriched whey protein was significantly associated with increased lower body function and muscle mass retention^123^. Our findings provide further evidence of the protective effects of leucine/isoleucine in PD and suggest them as possible interventional targets.

In the UKB, PGS for prioritised polygenic metabolites showed limited prediction of their respective outcome disorder. Polygenic scores are noted to explain only a small proportion of the variance even within the same phenotype^124^. As such, it is not entirely surprising that we are unable to detect significant associations cross-trait, particularly given the smaller effective target sample size available in the UKB compared to the large, meta-analytic samples sizes available from GWAS summary statistics for use in MR. To improve prediction, future studies might aggregate the genetic effects of multiple metabolites to better capture the total underlying metabolic risk for disease, for example through a multi-PGS framework^125^.

In conclusion, we performed metabolome-wide MR to test the causal effects of 1300 metabolites/metabolite ratios on eight neuropsychiatric disorders. Results implicate AA and LA-containing lipids across the psychiatric disorder spectrum, with shared causal variants identified near the *FADS* gene cluster. We also identified shared causal variants between glutamine metabolites and Alzheimer’s disease, and identify a shared effect of *SPRD4* gene expression. Additionally, our results implicate sphingolipid metabolism in psychiatric disorders and carnitine related metabolites as protective for MS and ALS, alongside several promising target metabolites with disease-specific effects. More broadly, this study suggests that lipid metabolism plays an important role in both psychiatric and neurodegenerative risk. The metabolites identified in this study can help inform potential interventions and future, targeted studies of metabolic contributions to disease risk.

## 5. Limitations

This study used data from individuals of European ancestry only and as such may lack generalisability. Further, as previously noted^79^, although the exclusion of the *APOE* region from the AD analyses is necessary to avoid violating MR assumptions, this may result in false negatives due to the regions known role in lipid metabolism^126^. Conversely, we can have greater confidence that the metabolites identified here are not confounded by *APOE* effects. Given the importance of this region in AD and lipid metabolism, it deserves specific focus in future metabolomic work. Further, MR requires that several assumptions – such the availability of suitable instruments and absence of pleiotropic effects – be met to provide reliable results. Although we mitigate against these by including only strong instruments (F-statistic ≥ 10) and using a robust sensitivity criterion to delineate causal metabolites, results should be interpreted with caution prior to further triangulation.

## 6. Funding and acknowledgements

LG is funded by the King’s College London DRIVE-Health Centre for Doctoral Training and the Perron Institute for Neurological and Translational. JM is supported by the King’s Prize Fellowship. This study has been partly delivered through the National Institute for Health and Care Research (NIHR) Maudsley Biomedical Research Centre (BRC). PP is funded by an Alzheimer’s Research UK Senior Research Fellowship.

This research has been conducted using the UK Biobank Resource under Application Number 82877. Ethical approval for the UK Biobank study has been granted by the National Information Governance Board for Health and Social Care and the NHS North West Multicentre Research Ethics Committee (11/NW/0382). Written informed consent was obtained from all participants by the UK Biobank. We thank the UK Biobank Team for collecting the data and making it available. We also thank the UK Biobank participants.

We also thank and acknowledge the contribution and use of the CREATE high-performance computing cluster at King’s College London (King’s College London. (2022). King’s Computational Research, Engineering and Technology Environment (CREATE)).

We want to acknowledge the participants of the FinnGen study and the veterans of the Million Veterans Project, as well as the research teams for both studies.

For the purposes of open access, the author has applied a Creative Commons Attribution (CC BY) license to any Accepted Author Manuscript version arising from this submission.

## 7. Data availability

Data is available on reasonable request from the UK Biobank (https://www.ukbiobank.ac.uk/learn-more-about-uk-biobank/contact-us).

Summary statistics from FinnGen are available online (https://www.finngen.fi/en/access_results) and from the Million Veterans Project via dbGaP request (https://www.ncbi.nlm.nih.gov/projects/gap/cgi-bin/study.cgi?studyid=phs001672.v1.p1).

GWAS summary statistics from the Psychiatric Genomics Consortium (PGC) for Alzheimer’s disease, anxiety, bipolar disorder, depression and schizophrenia are available from their website (https://pgc.unc.edu/for-researchers/download-results/). Summary statistics for anxiety from iPSYCH are also available online (https://ipsych.dk/en/research/downloads). For Parkinson’s disease and ALS, summary statistics are available on GWAS Catalog at https://www.ebi.ac.uk/gwas/studies/GCST009325 and https://www.ebi.ac.uk/gwas/studies/GCST90027164 respectively. For multiple sclerosis, summary statistics are available though the IEU Open GWAS Project (https://gwas.mrcieu.ac.uk/datasets/ieu-b-18/). Summary statistics for metabolites measured from Chen et al.^58^ are available on GWAS catalog (https://www.ebi.ac.uk/gwas/studies/) under accession numbers GCST90199621-90201020. Access to summary statistics for the metabolites measured by Hysi et al.^59^ were made available following request to the original authors.

## 8. Conflicts of Interest

CML is on the Scientific Advisory Board of Myriad Neuroscience and has received honoraria for consultancy from UCB. C.L.-Q. has received consultancy fees from Pfizer. C.L.-Q. has received honoraria, travel or speakers’ fees from Biogen and research funds from Pfizer and Novo Nordisk. C.L.-Q. is the director of the company BrainLogia. All other authors declare no competing interests.

## Notes

### Author Declarations

Ethics committee/IRB of King's College London gave ethical approval for this work Ethical approval for the UK Biobank study has been granted by the National Information Governance Board for Health and Social Care and the NHS North West Multicentre Research Ethics Committee (11/NW/0382). Data access permission has been granted under UK Biobank application 82877. Written informed consent was obtained from all participants by the UK Biobank.

