## Supplementary Materials for "Evaluating metabolome-wide causal effects on risk for psychiatric and neurodegenerative disorders"

**Supplementary Material 1: Description of genetic correlations between neuropsychiatric disorders**

Psychiatric disorders were strongly genetically correlated (*r_g_* range: 0.37-0.85, *p*-value range: 1.95 x 10^-148^ - 7.34 x 10^-66^, *p_FDR_* range: 5.46 x 10^-147^ - 3.43 x 10^-65^). This pattern was not observed between the neurodegenerative disorders, where only AD and ALS genetically correlated after FDR correction (*r_g_* [SE] = 0.34 [0.004], *p*-value = 6.75 x 10^-6^, *p_FDR_*  = 2.10 x 10^-5^). Between the psychiatric and neurodegenerative disorders, SCZ was significantly genetically correlated with AD (*r_g_* [SE] = 0.11 [0.003], *p*-value = 1.76 x 10^-3^; *p_FDR_* = 0.005). MS was genetically correlated with ANX, DEP and SCZ (*r_g_* range: 0.07- 0.13, *p*-value range: 1.48 10^-7^- 0.009, *p_FDR_* range: 5.90 x 10^-7^ – 0.02) (**Supplementary Material 2; Supplementary Table 1** and **2**).

**Supplementary Material 2: Heatmap of genetic correlations between neuropsychiatric disorders, calculated using linkage disequilibrium score regression (LDSC) within GenomicSEM.**

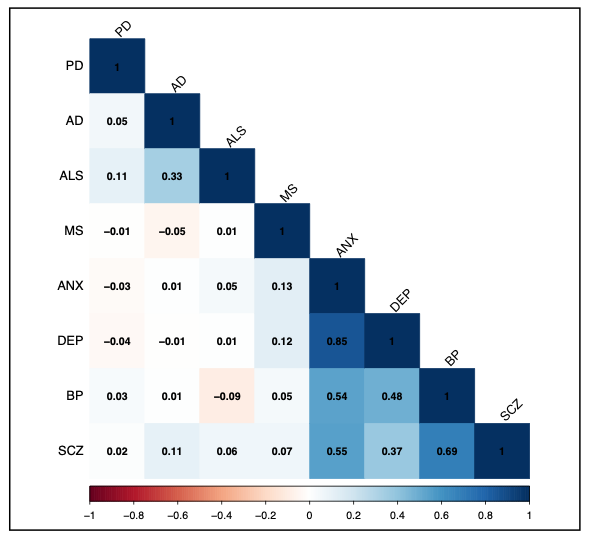

**Supplementary Material 3: Forest plots of IVW-MR estimates for the effect of significant polygenic metabolites on unadjusted and BMI, waist-to-hip ratio and educational attainment adjusted neuropsychiatric outcomes.**

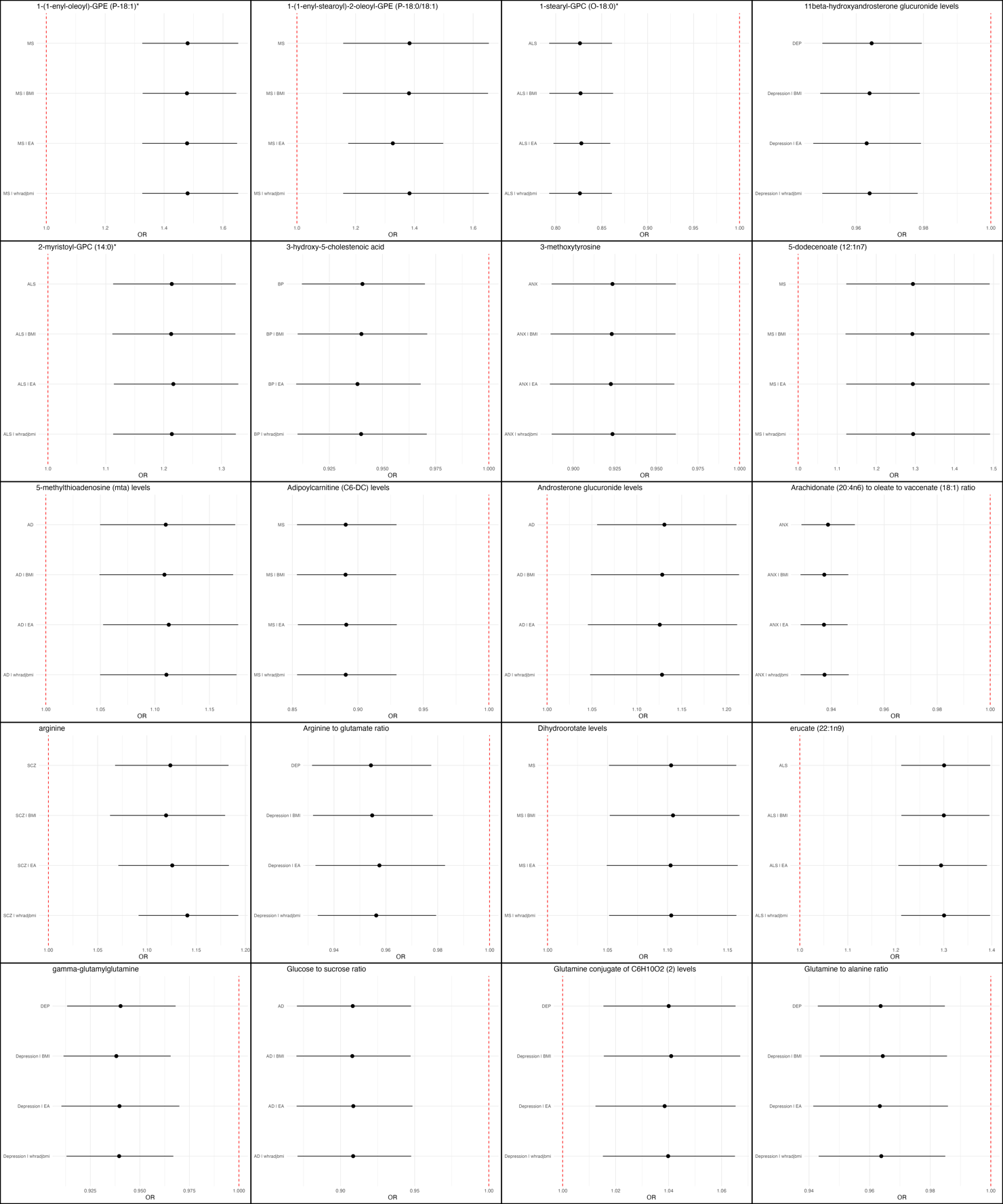

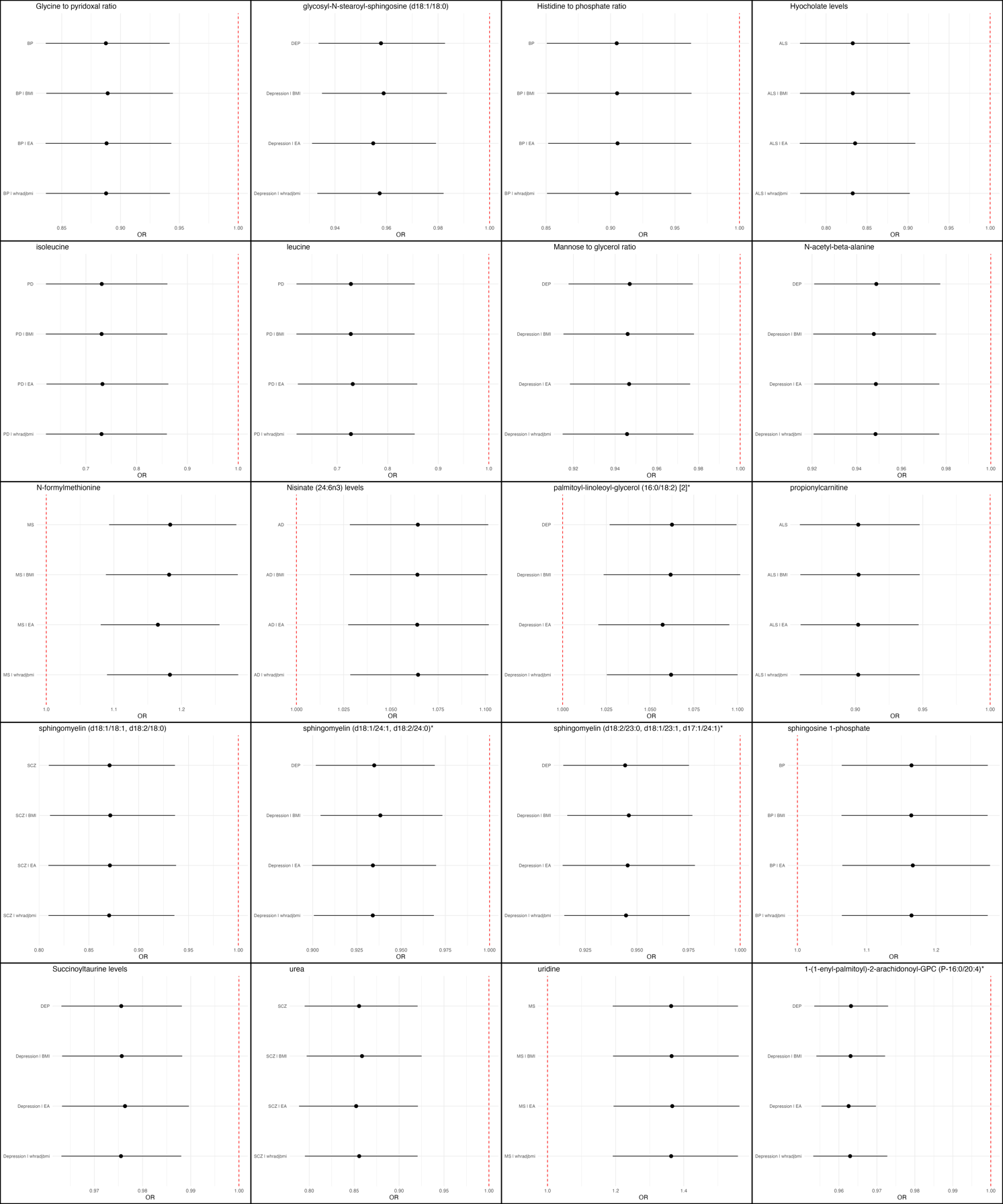

**Supplementary Material 4: Simplified metabolic pathway from linoleic acid (LA) to arachadonic acid (AA) derived from KEGG pathway hsa00591**

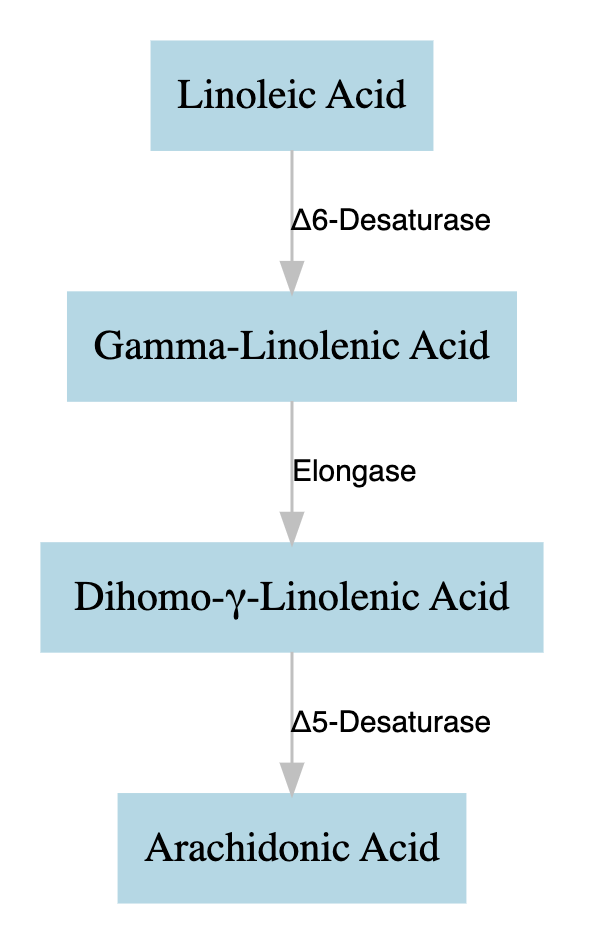

**Supplementary Material 5: Simplified metabolic pathway for sphingomyelin and ceramide metabolism from KEGG pathway hsa00600**

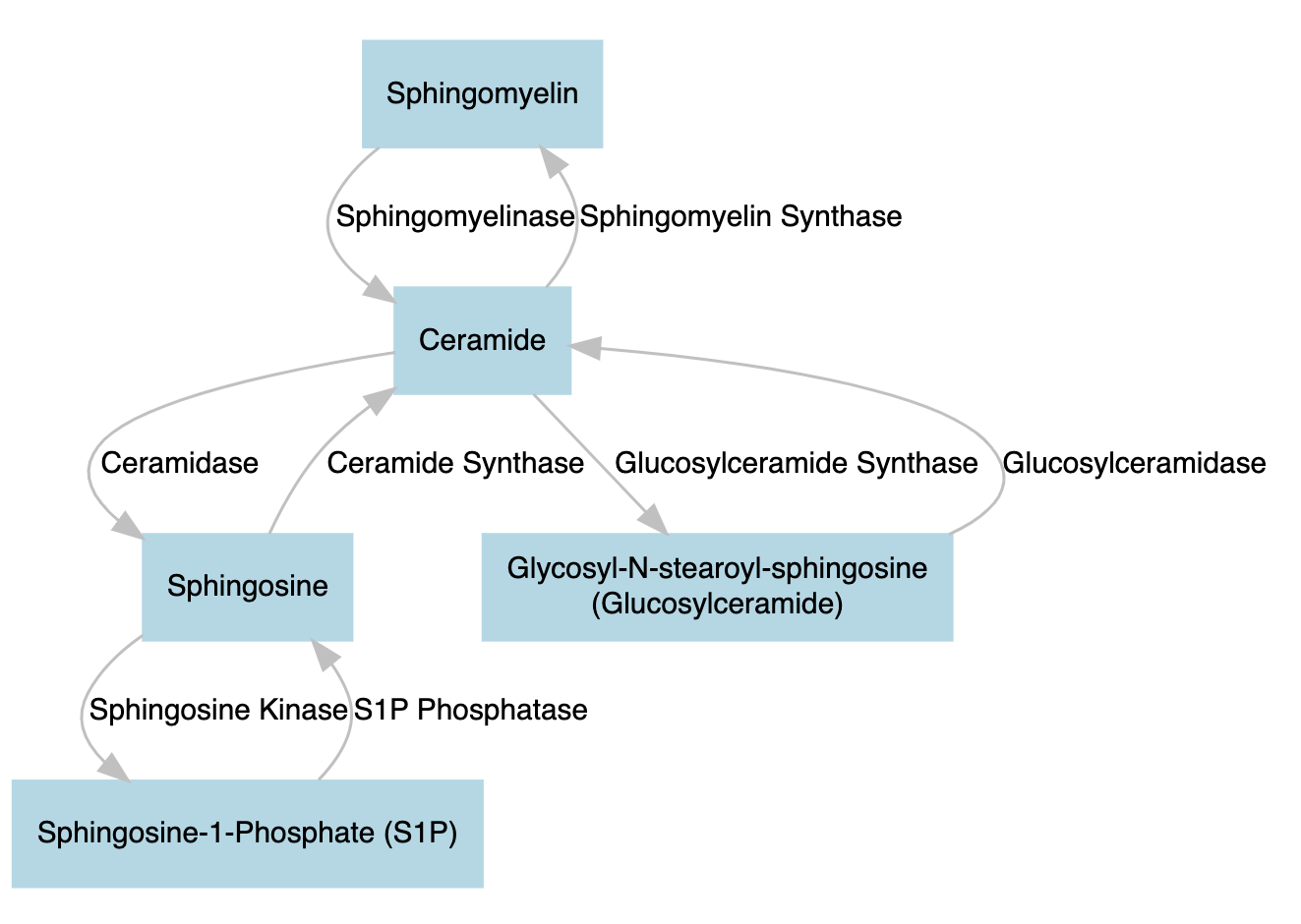
